# Value of 3D-echocardiography to improve prediction of mitral valve reoperation in patients with congenital mitral valve disease

**DOI:** 10.1101/2023.05.09.23289752

**Authors:** Nora Lang, Steven J. Staffa, David Zurakowski, Francesca Sperotto, Melinda Shea, Christopher W. Baird, Sitaram Emani, Pedro J. del Nido, Gerald R. Marx

**Affiliations:** Department of Cardiology, Boston Children’s Hospital, Harvard Medical School, Boston, Massachusetts, USA; Department of Surgery, Department of Anesthesiology, Critical Care and Pain Medicine, Boston Children’s Hospital, Harvard Medical School, Boston, Massachusetts, USA; Department of Cardiovascular Surgery, Boston Children’s Hospital, Harvard Medical School, Boston, Massachusetts, USA; Department of Pediatric Cardiology, University Heart & Vascular Center Hamburg, University Medical Center Hamburg-Eppendorf, Hamburg, Germany

**Keywords:** 3D-echocardiography, congenital mitral valve disease, mitral stenosis, mitral regurgitation, prediction, outcomes

## Abstract

**BACKGROUND:** Congenital mitral valve disease (CMVD) presents major challenges in medical and surgical management. Data on predictors of mitral valve (MV) reoperation and value of 3D echocardiography (3DE) in this context are currently limited. Aim of our study was to identify predictors of MV reoperation and to investigate the value of 3DE in risk stratification.

**METHODS:** All children <18 years old who underwent MV reconstruction for CMVD from 2002 to 2018 were included. Pre and postoperative 2D-echocardiogram (2DE) and 3DE data were collected. Competing risks and Cox regression analyses were used to identify independent predictors of MV reoperation. Receiver operating characteristics curve (ROC) analysis was used to assess the predictive values of 3DE versus 2DE. Decision tree analysis was used to compute decision algorithms.

**RESULTS:** Over the course of the study period, 206 children underwent MV reconstruction for CMVD: 105 had mitral stenosis (MS), 75 had mitral regurgitation (MR), and 26 had mixed disease (MD). Of them, 64 (31%) required a MV-reoperation. At multivariable analysis, age <1 year (hazard ratio [HR]=2.65; 95% confidence interval [CI]:1.13-6.21), tethered leaflets (HR=2.00; 95% CI:1.05- 3.82;), moderate or greater 2DE postoperative MR (HR=4.26; 95% CI:2.45-7.4), as well as changes in 3D-effective orifice area (3D-EOA) and in 3D-vena contracta regurgitant area (3D-VCRA) were all found to be independent predictors of MV reoperation. By ROC analysis, changes in 3D-EOA and 3D-VCRA were found to have significantly higher predictive value than changes in mean gradients (AUC=0.847 vs AUC=0.676, *P*=0.006) and 2D-VCRA (AUC=0.969 vs AUC=0.720, *P*=0.012), respectively. Decision-tree analysis found that a <30% increase in 3D-EOA had 80% accuracy (HR=8.50; 95% CI:2.9-25.1) and a <40% decrease in 3D-VCRA had 93% accuracy (HR=22.50; 95% CI:2.9-175) in predicting MV reoperation for stenotic and regurgitant MV, respectively.

**CONCLUSIONS:** Age <1 year, tethered leaflets, 2DE postoperative MR, changes in 3D-EOA and 3D-VCRA were all independent predictors of MV-reoperation. 3DE parameters had higher predictive value than 2DE. 3DE-based decision-tree algorithms may help risk stratification and serve as a support tool for clinical decision-making in patients with CMVD.

**CLINICAL PERSPECTIVES:** *What is new?:*

- In congenital mitral valve (MV) disease patients undergoing MV reconstruction, changes in 3D effective orifice area (3D-EOA) and 3D vena contracta regurgitant area (3D-VCRA) were found to be independent predictors of MV reoperation.
- Changes in 3D effective orifice area (3D-EOA) and 3D vena contracta regurgitant area (3D-VCRA) had significantly higher predictive value compared to 2D-echocardiography parameters, i.e. changes in mean gradient, 2D-VCRA and moderate or greater postoperative 2D mitral regurgitation.
- Decision tree analysis identified cut-off points in changes of 3D-EOA and 3D-VCRA to predict MV reoperation.

*What are the clinical implications?:*

- Changes in 3D-EOA and 3D-VCRA can be measured in the operating room to consider return to bypass, or at discharge, to help inform need for early follow-up and MV reoperation.
- 3DE-based decision tree predictive algorithms may help in risk stratification and serve as a support tool to facilitate clinical decision-making.

## INTRODUCTION

Congenital mitral valve disease (CMVD) is a rare and heterogenous congenital heart disease (CHD) with variable anatomic characteristics and prognosis. The disease may affect multiple segments of the valve apparatus including the supravalvar region, annulus, leaflets, commissures, as well as the subvalvar region^1–2^. Despite medical management, children with CMVD often require catheter-based or surgical interventions^3^. MV repair is generally preferred over MV replacement due to its ability to preserve the subvalvar apparatus and its function, conserve the overall ventricular geometry, and allow tissue growth over time^4–7^. However, studies have shown that surgical results have been burdened by a non-negligible proportion of reoperation for either MV reconstruction or replacement^3, 5, 8^.

The identification of factors possibly associated with higher risk of reoperation may help guide risk stratification and management in this peculiar cohort of patients. In the last decades, studies have tried to identify predictors of MV reoperation in patients with CMVD^5, 8–10^. However, often these studies were affected by small sample sizes, or included patients with marked heterogeneity in their baseline MV pathology^7, 9^. Few studies have addressed the value of echocardiography techniques to predict MV reinterventions^9–10^.

Traditionally, cardiologists and cardiac surgeons have relied on 2D-echocardiography (2DE) for monitoring and guiding the clinical management in these patients. 3D-echocardiography (3DE) provides the simultaneous assessment of the spatial relationship of the leaflets, chordae and papillary muscles, and thus more accurate and reliable measurements^11–13^, which have the potential to aid surgical planning^14^. To date, most reports assessing the benefit of 3DE in cardiac diseases have been conducted in adult patients^11, 15–16^. The use of 3DE in children with CHD has been reported only for specific settings, like the evaluation of atrioventricular (AV) septal defects *status post* repair ^17–18^ or the assessment of the tricuspid valve in children with hypoplastic left heart syndrome (HLHS) ^18–20^.

The main purpose of this study was to investigate independent predictors of MV reoperation in a large cohort of pediatric patients with CMVD. In addition to traditional patient demographics, anatomic and clinical characteristics, we sought to assess 3DE measurements as predictors of MV reoperation, and to investigate the relative performance of the 3DE compared to the 2DE in this setting. Finally, we aimed to develop decision-tree algorithms for patients’ risk stratification to serve as a support tool for clinicians in the clinical decision-making.

## MATERIAL AND METHODS

### Patients

The Cardiovascular Surgical Department database at Boston Children’s Hospital (Boston, MA) was searched for all patients <18 years who underwent MV surgery between January 2002 and December 2018. The Institutional Review Board at Boston Children’s Hospital approved the study with a waiver for informed consent (IRB-P00023266).

Patients with HLHS who underwent single ventricle palliation, patients with AV-canal defects, AV-discordance, and those with connective tissue disorders were excluded. Patients were subdivided into three subgroups according to the type of MV disease as follows: (1) mitral stenosis (MS) group: patients with a Doppler mean gradient >5 mmHg and none or trivial MR; (2) mitral regurgitation (MR) group: patients with a mean gradient <5 mmHg and at least moderate MR; (3) mixed disease (MD) group: patients with a mean gradient >5 mmHg and at least moderate MR. Anatomical characteristics of the MV were determined based on the description of the MV in the surgical report, as detailed in the **Supplemental Methods**.

### Outcomes

The primary outcome measure was MV reoperation. Secondary outcome measures were need for MV replacement and death at any time to last follow-up.

### Echocardiographic measurements

Echocardiographic measurements were performed preoperatively (within 7 days before surgery) and postoperatively either at hospital discharge or at 10 days after surgery, whichever came first.

### 2DE measurements

Qualitative grading of the MR was obtained from the echocardiographic report and was classified as trivial, mild, moderate, or severe. Doppler mean gradients and mitral 2D-VCRA were calculated by 2 independent investigators. The severity of MS was graded as follows: trivial: <3 mmHg; mild: 3–5 mmHg; moderate: >5 to 10 mmHg, and severe: >10 mmHg. 2D-VCRA was measured from two orthogonal diameters, and the area was calculated from the equation of an ellipse (π*(d/2)^2^).

### 3DE measurements

Electrocardiographic-gated full-volume 3DE acquisitions were performed using a 5–1 MHz matrix-array transthoracic probe and a 3DE ultrasound system (SONOS 7500 and iE33, Philips Medical Systems, Bothwell, WA). Full-volume 3DE data were acquired from the apical four chamber view. Analyses were performed with a dedicated software (Q-lab 6.0, Philips Medical Systems, Bothwell, WA). Multi-planar reconstruction was used to locate the cross-sectional plane to measure the annulus area, 3D-EOA and 3D-VCRA (**Figure 1A and B**). Based on two orthogonal long-axis planes, a corresponding short-axis plane was chosen to appropriately trace the MV annulus area, 3D-EOA, and 3D-VCRA (**Figure 1A and 1B**), which were blindly measured by 2 independent investigators. Each investigator repeated these measurements blindly after 2 weeks.

**Figure 1.**
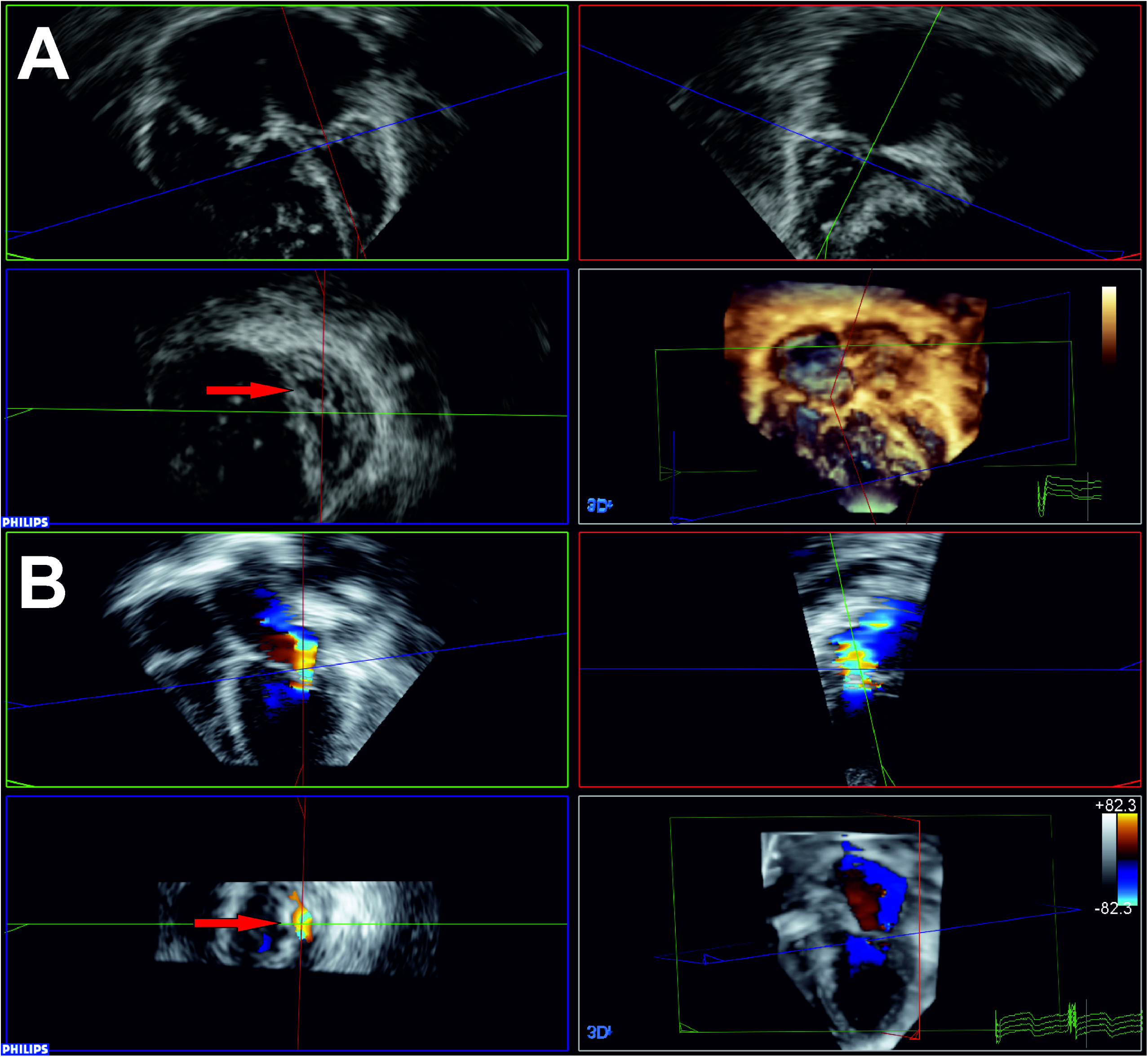
Measurement of 3D-EOA and 3D-VCRA. Multiplanar reconstruction was used to locate the optimal cross-sectional plane to determine the annulus area, 3D-EOA (**A**) and 3D-VCRA (**B**). The 3D-EOA was defined as the smallest orifice of the MV inflow. The 3D-VCRA corresponded to the cross-sectional area of the color Doppler jet at valve coaptation. **A.** For 3D-EOA measurement, the frame during diastolic opening with the largest opening was chosen and the MV was aligned in two long-axis orthogonal planes. A cross sectional plane (blue) was set at the desired position to allow for 3D-EOA circumference planimetry. **B.** For 3D-VCRA measurement, the longitudinal planes were adjusted to best visualize the regurgitant jet, allowing for placement of the cross-sectional plane at the largest circumference; 3D-VCRA was then measured before jet dispersion in orthogonal imaging planes. EOA: effective orifice area. MV: mitral valve. VCRA: vena contracta regurgitant area.

### Statistical Analysis

Demographic and anatomic characteristics are summarized using frequencies and percentages for categorical data, medians and interquartile ranges (IQR) for continuous data. Paired t-test was used to assess variation of echocardiographic parameters over time (pre and postoperatively).

The Kaplan Meier estimator was used to compute freedom from MV reoperation, MV replacement, and death at 1, 3, and 5 years after the first MV operation and 95% confidence intervals (CIs). Inter and intra-rater agreement of 3DE measurements were tested using intra class correlation coefficients (ICC) based on a two-way mixed effects modeling, with reliability categories defined as follows: ICC <0.50 poor, 0.50-0.75 moderate, 0.75-0.90 good, >0.90 excellent.

Univariate and multivariable competing risk regression analyses using Fine-Gray modeling were used to identify significant demographic, clinical, and 2DE predictors of MV reoperation, accounting for mortality prior to reoperation as a competing risk event^21–22^. Preoperative factors included in the model were age, weight, sex, type of disease (MS, MR, MD), pulmonary hypertension, thickened and tethered leaflets, presence of endocardial fibroelastosis (EFE), year of surgery; postoperative factors were presence of moderate or greater MR and presence of moderate or greater MS postoperatively. All factors were included in the multivariable model. Cumulative incidence functions were constructed overall and for diagnostic subgroups for independent predictors using Nelson-Aalen estimators. To better investigate any independent 2DE predictor resulted from this model, the same model was reproduced for the subgroups of patients with MS (including MD) and MR (including MD). 3DE measurements were tested as predictors of MV-reoperation using univariate and multivariable time-to-event Cox proportional hazards regression analysis, for the MS (including MD) and the MR (including MD) populations separately. Factors found to be significant at univariate analysis were included in the multivariable model. The proportional sub-distribution hazard assumption was tested using Schoenfeld residuals ^23^ and the Grambsch-Therneau test. Results are presented as adjusted hazard ratios (HR) and 95% CIs.

The prognostic value of 2DE versus 3DE measurements in predicting MV-reoperation was assessed by the area under the curve (AUC) of the receiver operating characteristic curves, which were compared using the paired-DeLong test^24^. Classification and regression tree analysis was implemented to determine the optimal cut-points for 3DE measurements in predicting MV reoperation (*rpart* package, R). Results from decision-tree analysis are presented with sensitivity, specificity, positive predictive value (PPV), negative predictive value (NPV), and accuracy of the optimized predictive cutoff thresholds. Bootstrap validation was used to evaluate the internal validity and model performance^25^ (**Supplemental Methods**). Statistical analyses were performed using Stata (version 16.0, StataCorp LLC, College Station, Texas) and R (version 3.4.3, R Foundation for Statistical Computing, Vienna, Austria). A two-tailed alpha level of 0.05 was considered statistically significant.

## RESULTS

### Demographic and anatomic MV characteristics

Two-hundred-and-six patients (48% female) underwent MV surgery during the study period. One-hundred-and-five patients (51%) had MS, 75 (36%) MR, and 26 (13%) MD. Eight percent were neonates, 47% were infants. The median age at the initial MV operation was 17 months (IQR: 5-56), the median weight was 9.1 kg (IQR: 5-16.7). Surgical anatomic MV characteristics are shown in **Table 1**. At time of the initial operation, MD patients were younger (median age 5 months, IQR: 1-21) and smaller in weight (median weight 5.1 kg, IQR: 3.3–11.4). MR patients were older (median age 48 months, IQR: 5–73) and weighed more (median weight 13.7 kg, IQR: 5.2-19.6). Median follow-up for the total cohort was 60 months (IQR: 18-108) (**Table 1**).

**Table 1.** Patients’ baseline demographic and anatomical characteristics.

|  | **Total Cohort**<br>**(n=206)** | **Mitral Stenosis**<br>**(n=105)** | **Mitral**<br>**Regurgitation**<br>**(n=75)** | **Mixed Disease**<br>**(n=26)** |
| --- | --- | --- | --- | --- |
| **Demographics** |  |  |  |  |
| **Median age, months (IQR)** | 17.3 (4.6 - 56) | 11.9 (5.3 - 43.7) | 47.7 (5.3 – 72.9) | 4.6 (1.4 – 21.2) |
| **Median weight, kg (IQR)** | 9.1 (5 – 16.7) | 8.4 (5.4 – 13.2) | 13.7 (5.2 – 19.6) | 5.1 (3.3 – 11.4) |
| **Median follow up, months (IQR)** | 59.4 (18.3- 108.4) | 63 (15.3 – 115.7) | 55.2 (16.8 – 87.4) | 76.8 (27.4 -108.4) |
| **Anatomical characteristics** |  |  |  |  |
| **Double orifice mitral valve** | 13 (6.3%) | 8 (7.6%) | 3 (4%) | 2 (7.7%) |
| **Annular dilatation** | 50 (24.3%) | 4(3.8%) | 39 (52%) | 7 (26.9%) |
| **Cleft leaflet** | 46 (22.3%) | 0 (0%) | 39 (52%) | 7 (26.9%) |
| **Elongated chordae** | 10 (4.9%) | 0 (0%) | 10 (13.3%) | 0 (0%) |
| **Shortened chordae** | 76 (36.9%) | 47 (44.8%) | 17 (22.7%) | 12 (46.2%) |
| **Absent chordae** | 14 (6.8%) | 6 (5.7%) | 4 (5.3%) | 4 (15.4%) |
| **Fused/closely spaced chordae** | 45 (21.8%) | 37 (35.2%) | 3 (4%) | 5 (19.2%) |
| **Secondary/abnormal chordae** | 84 (41.0%) | 39 (37.1%) | 26 (34.7%) | 17 (65.4%) |
| **Commissural fusion** | 30 (14.6%) | 26 (24.8%) | 0 (0%) | 4 (15.4%) |
| **Hammock valve (mitral arcade)** | 22 (10.7%) | 12 (11.4%) | 3 (4%) | 7 (26.9%) |
| **Stenosing mitral membrane** | 65 (31.6%) | 62 (59.0%) | 0 (0%) | 3 (11.5%) |
| **Endocardial fibroelastosis** | 62 (30.1%) | 46 (43.8%) | 8 (10.7%) | 8 (30.8%) |
| **Single dominant papillary muscle** | 27 (13.1%) | 22 (21.0%) | 4 (4%) | 1 (3.8%) |
| **Prolapse of the anterior leaflet** | 51 (24.8%) | 5 (4.8%) | 35 (46.7%) | 11 (42.3%) |
| **Thickened leaflets** | 110 (53.4%) | 64 (61.0%) | 30 (40.0%) | 16 (61.5%) |
| **Tethered leaflets** | 115 (55.8%) | 58 (55.2%) | 34 (45.3%) | 23 (88.5%) |
| **Tethered papillary muscles** | 94 (45.6%) | 51 (48.6%) | 23 (30.7%) | 20(76.9%) |
| **Closely spaced papillary muscles** | 34 (16.5%) | 28 (26.7%) | 0 (0%) | 6(23.1%) |

### Mortality

At a median follow-up of 60 months (IQR: 18-108), 13 (6%) patients did not survive. Death occurred at a median of 15 months (IQR: 2-42) after the first MV operation. The highest mortality rate was observed in MD patients (4/26, 15%), while the lowest was reported in MR patients (2/75, 3%). There was one peri-operative/early death (<30 days) in 2005. The overall survival was 97.2% at 1 year (95% CI: 93.3%-98.8%; n=164), 94.7% at 3 years (95% CI: 90%-97.2%; n=131) and 93.9% at 5 years (95% CI: 88.9%-96.7%; n=98).

### MV Reoperation and replacement

Sixty-four patients (31%) required MV reoperation at a median time of 9 months after the first operation (IQR: 0.4-39). Thirty-three (16%) patients required more than one MV reoperation. Twenty-six (12.6%) required a MV replacement, at a median time of 46 months since the first operation (IQR: 0.9-18). Overall freedom from MV reoperation was 79.1% at 1 year (95% CI: 72.4%-84.4%; n=132), 73.9% at 3 years (95% CI: 66.6%-79.8%; n=97) and 66% at 5 years (95% CI: 57.7%-73%; n=64). Thirty-six patients who underwent reoperation had MS (34% of MS), 16 MR (21% of MR), and 12 MD (46% of MD). Overall freedom from MV-replacement was 89.9% at 1 year (95% CI: 84.4%-93.5%; n= 150), 87.9% at 3 years (95% CI: 82%-91.9%; n=114) and 85.4% at 5 years (95% CI: 78.9%-90%; n=82). Seventeen patients who underwent MV replacement had MS (16% of MS), 4 MR (5% of MR), and 5 MD (19% of MD).

### Demographic, anatomic and 2DE predictors for MV-reoperation

At univariate competing risks regression analysis, age <1 year, MD, tethered leaflets, and moderate or greater postoperative MR or MS were found to be associated with MV reoperation (**Table 2**). Multivariable modeling (**Table 2**) confirmed that age <1 year (adjusted HR=2.65; 95% CI: 1.13-6.21), tethered leaflets (adjusted HR=2.00; 95% CI: 1.05-3.82) and a qualitatively moderate or greater postoperative MR (adjusted HR=4.26; 95% CI: 2.45-7.40; **Figure 2A**) were independent predictors of MV reoperation. When stratifying for type of MV disease, moderate or greater postoperative MR was confirmed to be an independent predictor for both patients with stenotic MV (**Figure 2B**; adjusted HR=3.21; 95% CI: 1.73-6.00) and patients with regurgitant MV (**Figure 2C**; adjusted HR=7.38; 95% CI: 3.46-15.70).

**Figure 2.**
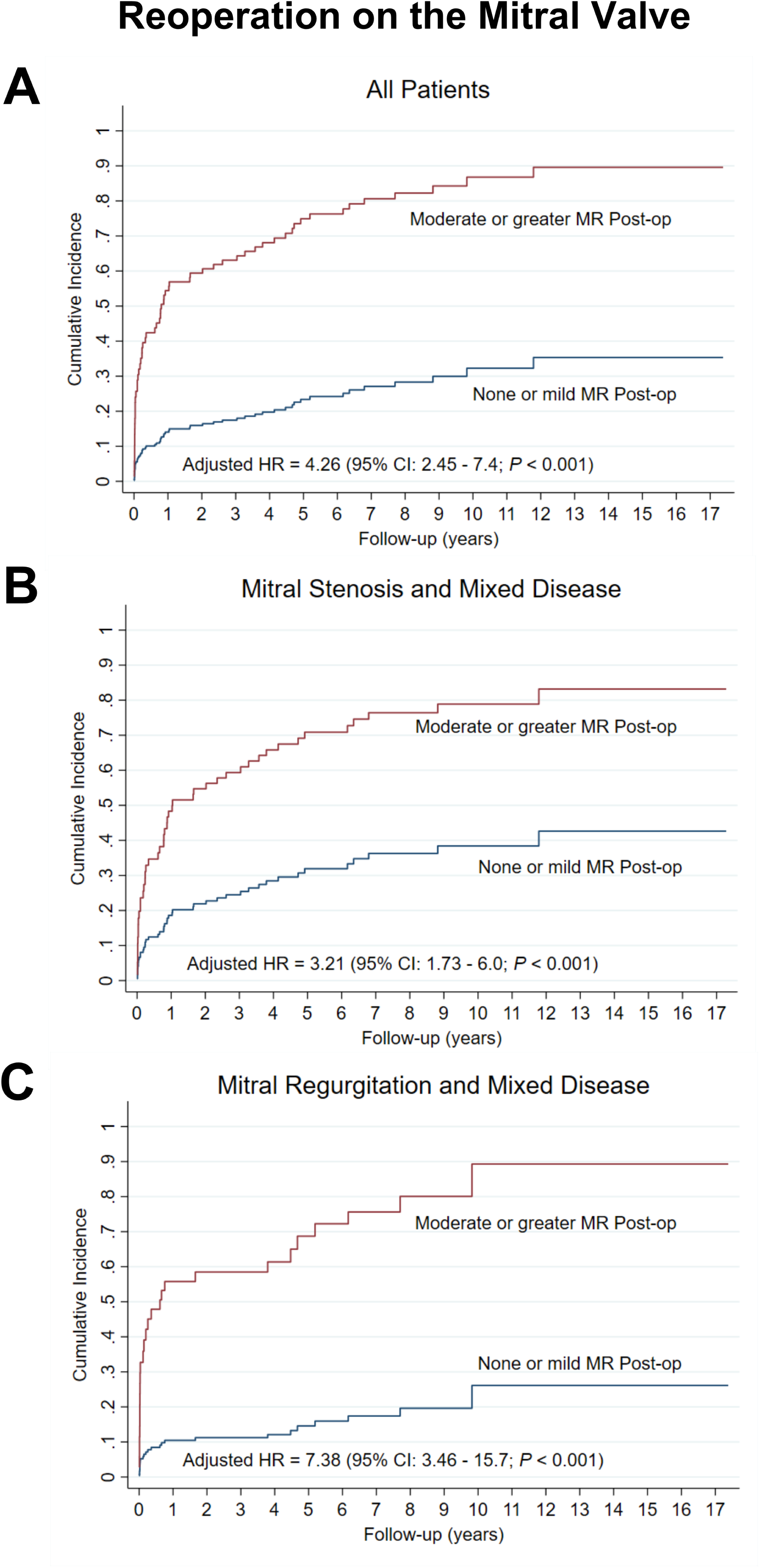
Competing risk analysis cumulative incidence curves for MV-reoperation according to evidence of moderate or grater postoperative MR by 2DE. **A.** The risk of MV reoperation was significantly higher in patients with moderate or greater postoperative MR (adjusted HR=4.26; 95% CI: 2.45-7.4). **B,C**. This association persisted for each baseline anatomic disease group (MS+MD patients: adjusted HR=3.21; 95% CI: 1.73-6.0; MR+MD patients: adjusted HR=7.38; 95% CI: 3.46-15.7). Multivariable models were adjusted for demographic, anatomic, and clinical characteristics shown in **Table 2**. 2DE: 2D echocardiography, EOA: effective orifice area, HR: hazard ratio, CI: confidence interval, MD: mixed disease, MR: mitral regurgitation, MS: mitral stenosis, MV: mitral valve, VCRA: vena contracta regurgitant area.

**Table 2.** Univariate and Multivariable Competing Risks Analysis to identify predictors of mitral valve reoperation

| Variable | Univariate Analysis |  |  |  | Multivariable Analysis |  |  |
| --- | --- | --- | --- | --- | --- | --- | --- |
|  | N | HR | 95% CI | P Value | Adjusted HR | 95% CI | P Value |
| **Baseline and preoperative characteristics** |  |  |  |  |  |  |  |
| Age < 1 year | 200 | 3.26 | (1.92, 5.52) | **<0.001\*** | 2.65 | (1.13, 6.21) | **0.025\*** |
| Weight (kg) | 206 | 0.96 | (0.91, 1.01) | 0.076 | 0.99 | (0.96, 1.04) | 0.726 |
| Biological gender | 195 |  |  |  |  |  |  |
| Female |  | Reference | . | . | Reference | . | . |
| Male |  | 1.18 | (0.71, 1.95) | 0.524 | 1.21 | (0.70, 2.10) | 0.494 |
| Type of disease | 206 |  |  |  |  |  |  |
| Mitral stenosis |  | 1.71 | (0.94, 3.14) | 0.081 | 1.13 | (0.56, 2.28) | 0.726 |
| Mitral regurgitation |  | Reference | . | . | Reference | . | . |
| Mixed |  | 2.57 | (1.16, 5.71) | **0.020\*** | 1.29 | (0.50, 3.31) | 0.600 |
| Pulmonary hypertension | 195 | 1.43 | (0.87, 2.35) | 0.159 | 0.87 | (0.49, 1.56) | 0.646 |
| Thickened leaflets | 206 | 1.22 | (0.74, 2.00) | 0.435 | 1.2 | (0.64, 2.27) | 0.570 |
| Tethered leaflets | 206 | 2.11 | (1.23, 3.61) | **0.007\*** | 2 | (1.05, 3.82) | **0.035\*** |
| Endocardial fibroelastosis | 205 | 1.16 | (0.69, 1.95) | 0.583 | 0.71 | (0.40, 1.29) | 0.263 |
| Year of surgery | 206 | 1.02 | (0.96, 1.09) | 0.472 | 1.03 | (0.95, 1.12) | 0.443 |
| **Postoperative imaging on 2D Echo** |  |  |  |  |  |  |  |
| 2D Moderate or greater MR Postop | 193 | 5.2 | (3.07, 8.80) | **<0.001\*** | 4.26 | (2.45, 7.40) | **<0.001\*** |
| 2D Moderate or greater MS Postop | 201 | 2.03 | (1.18, 3.46) | **0.010\*** | 1.67 | (0.94, 2.97) | 0.082 |

Competing risks modeling was computed using the Fine and Gray model, with mortality prior to reoperation as the competing event.
All the factors assessed at the univariate analysis (left) were included in the multivariable model. Multivariable model: N=183 patients,
57 of whom underwent MV-reoperation. MR: mitral regurgitation. MS: mitral stenosis. \*Statistically significant.

### Inter and intra-rater agreement of 3DE measurements

The inter-rater agreement was good for 3D-EOA (ICC=0.75), and excellent for both the 3D-VCRA and annulus area (ICC=0.93 and ICC=0.98, respectively). Intra-rater reliability was excellent for all parameters (EAO: ICC=0.92; 3D-VCRA: ICC=0.97; annulus area: ICC=0.99).

### 3DE predictors for MV reoperation

**Supplementary Video 1 and 2** show examples of 3DE in two patients with congenital MS and MR, respectively. Eighty-six out of 206 patients (42%) had pre and postoperative 3DE data available. For stenotic valves, the 3D-EOA significantly increased after MV-reoperation, from 0.91±0.55 cm^2^/m^2^ to 1.44±0.69 cm^2^/m^2^ (*P*<0.001). 3D-VCRA trended to increase from 0.93±1.50 cm^2^/m^2^ to 1.49±2.07 cm^2^/m^2^, although not significantly (*P=0.143*). On univariate analysis, preoperative 3D-EOA, early changes in 3D-EOA and 3D-VCRA, and size of the postoperative 3D-VCRA were positively associated with MV reoperation. On multivariable analysis, early changes in 3D-EOA and 3D-VCRA were confirmed to be independently predictive of MV reoperation (**Table 3**; 3D-EOA: adjusted HR=0.26; 95% CI: 0.07-0.99; and 3D-VCRA: adjusted HR=2.86; 95% CI: 1.09-7.48). For regurgitant valves, the 3D-VCRA significantly decreased from 2.22±2.26 cm^2^/m^2^ to 1.46±1.94 cm^2^/m^2^ (*P*=0.017), 3D-EOA significantly decreased from 2.91±1.86 cm^2^/m^2^ to 1.63±0.73 cm^2^/m^2^ (*P=0.005*), and the annuli significantly decreased from 7.53±3.45 cm^2^/m^2^ to 4.53±1.85 cm^2^/m^2^ (*P*<0.001). Univariate analysis showed that the size of the preoperative 3D-VCRA, size of the postoperative 3D-VCRA, early changes in 3D-VCRA, and preoperative annulus were positively associated with MV reoperation. On multivariable analysis, early changes in 3D-VCRA were confirmed to be an independent predictor of MV reoperation (**Table 3**; adjusted-HR=11.77; 95% CI: 3.06-45.3).

**Table 3.** Univariate and multivariable Cox regression analysis to predict mitral valve reoperation using 3D Echo variables

| **Mitral Stenosis and Mixed Disease Patients** |  |  |  |  |  |  |  |
| --- | --- | --- | --- | --- | --- | --- | --- |
| **3D Echo Variable** | **N** | **Univariate Cox Analysis** |  |  | **Multivariable Cox Analysis** |  |  |
|  |  | **HR** | **95% CI** | **P Value** | **Adjusted HR** | **95% CI** | **P Value** |
| Size of 3D-EOA preoperatively | 50 | 2.79 | (1.48, 5.25) | **0.002\*** | 1.30 | (0.56, 3.00) | 0.539 |
| Size of 3D-EOA postoperatively | 49 | 0.69 | (0.35, 1.36) | 0.283 |  |  |  |
| Change of the 3D-EOA | 39 | 0.27 | (0.12, 0.59) | **0.001\*** | 0.26 | (0.07, 0.99) | **0.049\*** |
| Size of 3D-VCRA postoperatively | 25 | 1.42 | (1.06, 1.90) | **0.018\*** | 0.75 | (0.37, 1.53) | 0.430 |
| Change of 3D-VCRA | 18 | 2.52 | (1.16, 5.48) | **0.020\*** | 2.86 | (1.09, 7.48) | **0.032\*** |
| **Mitral Regurgitation and Mixed Disease Patients** |  |  |  |  |  |  |  |
| **3D Echo Variable** | **N** | **Univariate Cox Analysis** |  |  | **Multivariable Cox Analysis** |  |  |
|  |  | **HR** | **95% CI** | **P Value** | **Adjusted HR** | **95% CI** | **P Value** |
| Size of 3D-EOA postoperatively | 30 | 0.80 | (0.39, 1.64) | 0.536 |  |  |  |
| Change of 3D-EOA | 30 | 0.96 | (0.32, 2.82) | 0.930 |  |  |  |
| Size of 3D-VCRA preoperatively | 38 | 1.81 | (1.19, 2.74) | **0.005\*** | 1.86 | (0.84, 4.11) | 0.127 |
| Size of 3D-VCRA postoperatively | 30 | 3.15 | (1.86, 5.36) | **<0.001\*** | 1.05 | (0.38, 2.91) | 0.921 |
| Change of 3D-VCRA | 30 | 15.7 | (4.65, 53.00) | **<0.001\*** | 11.77 | (3.06, 45.3) | **<0.001\*** |
| Size of the 3D annulus preoperatively | 45 | 1.03 | (0.91, 1.17) | 0.649 |  |  |  |
| Size of the 3D annulus postoperatively | 30 | 1.16 | (0.98, 1.39) | 0.086 |  |  |  |
| Change in 3D annulus size | 29 | 4.24 | (1.00, 18.3) | **0.050\*** | 1.93 | (0.28, 13.4) | 0.506 |

Factors found to be statistically significant at univariate analysis were included in the multivariable model. Multivariable analysis for patients with mitral stenosis and mixed disease: N=18. Multivariable analysis for patients with mitral regurgitation and mixed disease: N=25. 3D-EOA: 3D effective orifice area; 3D-VCRA: 3D vena contracta regurgitant area. \*Statistically significant.

### Comparison of 2DE vs 3DE measurements predictive values

**Figure 2** and **Table 4** showed the comparison of predictive values of 3DE vs 2DE parameters. When considering stenotic valves, early changes in 3D-EOA was found to be a stronger predictor of MV reoperation than 2DE early changes in mean gradients (AUC 0.847 [95% CI: 0.723-0.970] vs 0.676 [95% CI: 0.508-0.844], respectively; DeLong test *P*=0.006, **Figure 3A**). For regurgitant valves, early changes in 3D-VCRA was found to be a stronger predictor of MV reoperation than 2DE qualitative postoperative MR (AUC=0.969 [95% CI: 0.916-0.999] vs 0.751 [95% CI: 0.642-0.860], respectively; DeLong test *P<0.001*). Additionally, early changes in 3D-VCRA was a significantly stronger predictor of MV-reoperation than early changes in 2D-VCRA (AUC=0.969 [95% CI: 0.916-0.999] vs 0.720 [95% CI: 0.424-0.903], DeLong test *P*=0.012; Figure 2B).

**Figure 3.**
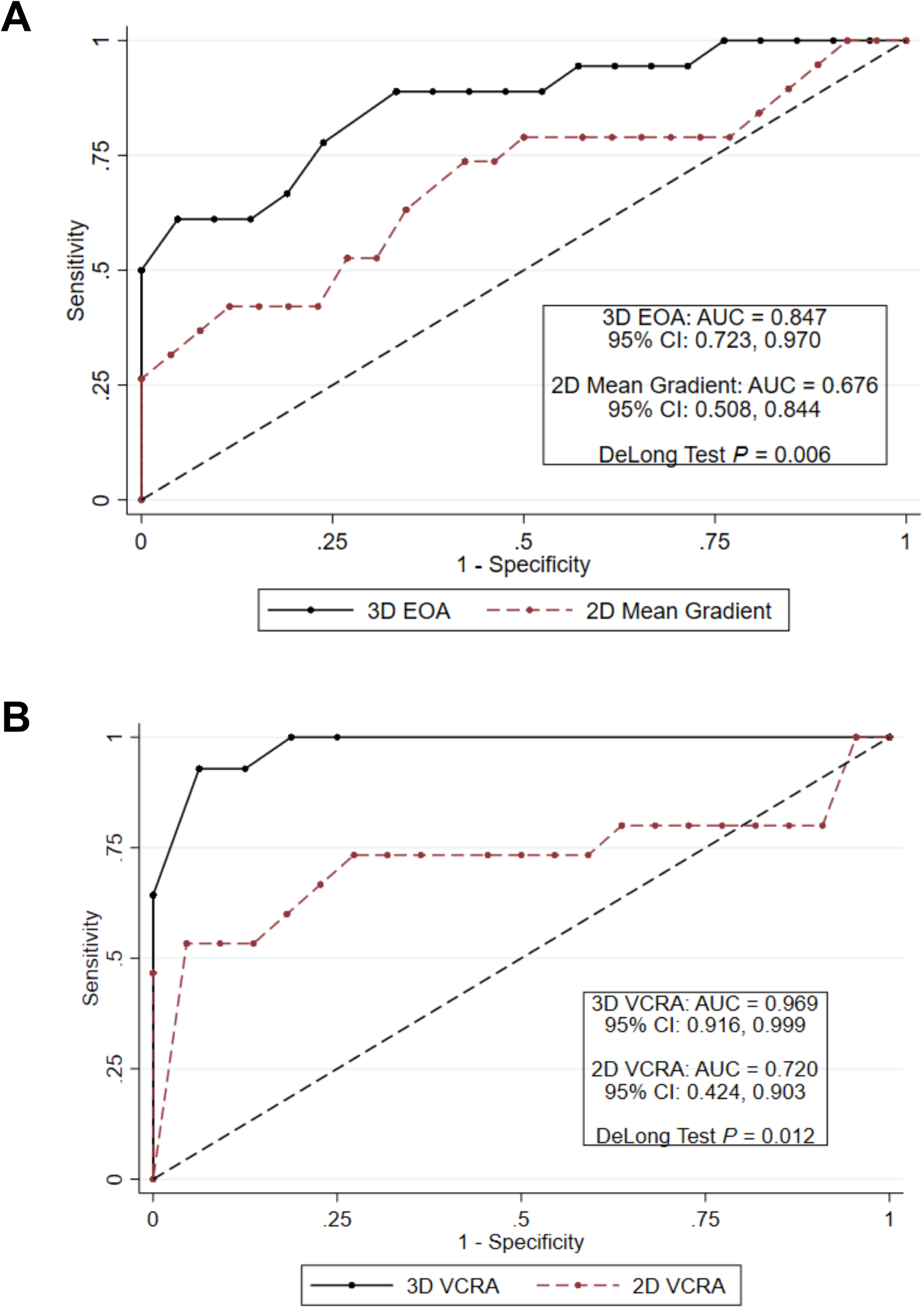
Comparison of 2DE versus 3DE predictive accuracy in predicting MV reoperation by AUC. **A.** When considering stenotic valves, AUC curve comparison revealed that early changes in 3D-EOA was a stronger predictor of MV reoperation than early changes in 2DE mean gradients (AUC 0.847 [95% CI: 0.723, 0.970] vs 0.676 [95% CI: 0.508, 0.844], respectively; DeLong test *P*=0.006). **B.** For regurgitant valves, AUC analysis revealed that early changes in 3D-VCRA was a significantly stronger predictor of MV reoperation than 2DE early changes in 2D-VCRA (AUC=0.969 [95% CI: 0.916, 0.999] vs 0.720 [95% CI: 0.424, 0.903], DeLong test *P*=0.012). 2DE/3DE: 2D/3D echocardiography, AUC: area under the curve, EOA: effective orifice area, CI: confidence interval, MV: mitral valve, VCRA: vena contracta regurgitant area.

**Table 4.** Predictive value of 2D and 3D echocardiography in assessing the risk of mitral valve reoperation based on receiver operating characteristics curve analysis and decision tree algorithms

| **Mitral Stenosis and Mixed Disease Patients** |  |  |
| --- | --- | --- |
| **Metric** | **Change in 2D**<br>**mean gradient** | **Change in**<br>**3D-EOA** |
| AUC (95% confidence interval) | 0.676 (0.508, 0.844) | 0.847 (0.723, 0.970) |
| Best cut-off identified by the decision tree<br>algorithm | < 30% increase | < 30% increase |
| Sensitivity | 88% (23/26) | 61% (11/18) |
| Specificity | 37% (7/19) | 95% (20/21) |
| PPV | 66% | 92% |
| NPV | 70% | 74% |
| **Mitral Regurgitation and Mixed Disease Patients** |  |  |
| **Metric** | **Change in**<br>**2D-VCRA** | **Change in**<br>**3D-VCRA** |
| AUC (95% confidence interval) | 0.720 (0.424, 0.903) | 0.969 (0.916, 0.999) |
| Best cut-off identified by the decision tree<br>algorithm | < 40% decrease | < 40% decrease |
| Sensitivity | 47% (7/15) | 93% (13/14) |
| Specificity | 95% (21/22) | 94% (15/16) |
| PPV | 88% | 93% |
| NPV | 72% | 94% |

### Decision-tree predictive algorithms for risk stratification

Decision-tree predictive algorithms were computed as a support tool for clinical decision-making (**Figure 4**). Sensitivity, specificity, positive and negative predictive values are reported in **Table 4**. For stenotic valves, an increase in 3D-EOA by <30% leads to 92% risk of MV-reoperation (accuracy 80%; HR=8.50; 95% CI: 2.90-25.1; *P*<0.001). For regurgitant valves, a decrease in 3D-VCRA <40% is associated with 93% risk of MV reoperation (accuracy 93%; HR=22.50; 95% CI: 2.90-175.00; *P*=0.003). Bootstrap validation demonstrated excellent internal validity and model performance (**Supplemental Results**).

**Figure 4.**
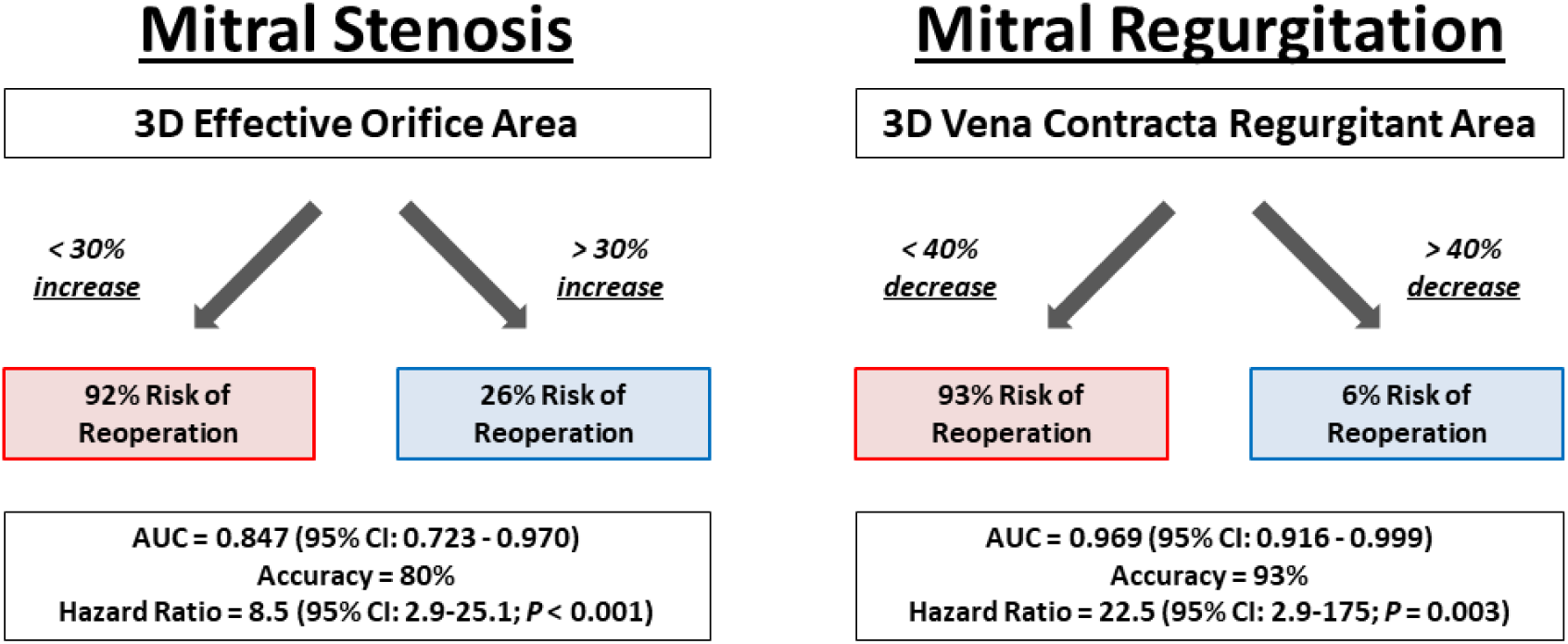
Decision-tree predictive algorithms for risk stratification. For stenotic valves, an increase in 3D-EOA by <30% leads to 92% risk of MV reoperation (accuracy 80%; HR=8.5; 95% CI: 2.9-25.1). For regurgitant valves, a decrease in 3D-VCRA <40% is associated with 93% risk of reoperation (accuracy 93%; HR=22.5; 95% CI: 2.9-175). Sensitivity, specificity, PPV and NPV are reported in **Table 4**. Bootstrap validation demonstrated excellent internal validity and model performance (**Supplemental Results**). AUC: area under the curve, EOA: effective orifice area, CI: confidence interval, hazard ratio: HR, MV: mitral valve, NPV: negative predictive value, PPV: positive predictive value, VCRA: vena contracta regurgitant area.

## DISCUSSION

This single-center study, which involved a large cohort of young pediatric patients undergoing MV surgery for CMVD, showed a low mortality at a median follow-up of 5 years. However, 31% of patients required a MV reoperation, and 12% requiring a subsequent MV replacement. These important surgical findings prompted analysis of risk factors for MV reoperation. We showed that age <1 year, presence of tethered leaflets, evidence of moderate or greater postoperative MR on 2DE, as well as early changes in 3D-EOA and 3D-VCRA were all independent predictors of MV reoperation. Importantly, when 2DE and 3DE were compared in terms of their performance to predict MV reoperation, certain 3DE parameters were found to have significantly higher prognostic values compared to 2DE. Based on these findings, decision tree predictive algorithms were developed to inform patients risk stratification, with the aim to improve assessment and help clinical decision-making.

The fact that younger patients, in particular patients <1 year of age, are at increased risk for MV reoperation was previously reported for smaller cohorts^5, 7–8, 10, 26^, and further confirmed in our study. Additionally, in terms of baseline MV morphology, our study showed that patients with tethered and restricted leaflets are at increased risk of MV reoperation. This is a new finding compared to what was previously reported in literature. From an echocardiographic point of view, we found that both 2DE and 3DE parameters were associated with MV reoperation. In particular, evidence of moderate or greater postoperative MR on 2DE, as well as early changes in the 3D-EOA and 3D-VCRA, were all found to be independently associated with the need for MV reoperation.

Postoperative systemic AV-valve regurgitation has been shown to be associated with adverse outcomes in a variety of CHDs^27–30^. In 43 adults with CHD undergoing primary or re-operative systemic AV-valve surgery, pre-discharge systemic AV-valve regurgitation grade was the only factor associated with adverse outcomes including reoperation ^27^. Similarly, studies have shown that postoperative systemic AV-valve regurgitation in repaired AV-canal is associated with higher risk of AV-valve reoperation^28–30^. Given the proven importance of this factor in determining outcomes, studies have also focused on improving 2DE regurgitation assessment with the evaluation of other parameters, such as 2D-VCRA. However, Prakash et al. did not find that 2D-VCRA measurements in patients with AV septal defects was superior to the qualitative regurgitation assessment^31^, and Yosefy et al. showed that 2D-VCRA can cause clinical misclassification in 45% of adult patients with eccentric MR, while 3D-VCRA was more accurate ^32^. In fact, non-negligible changes in the AV-valve geometry have been demonstrated in similar populations with CHD after AV-valve surgery, as in children with AV-canal undergoing AV-valve repair^33^. To date, measurements of 2D-VCRA are performed by measuring the vena contracta width mostly in one plane, and in some studies in two orthogonal planes. However, these types of measurement do not account for the marked irregularity of shape of the VCRA.

Several publications in adults have highlighted the advantages of 3DE for the quantitative assessment of the valve function^34–38^, with increasing number of reports demonstrating higher accuracy of 3DE compared to 2DE, and higher ability to predict outcome^39–40^. In parallel, a new consensus document recently advised on the use of 3DE for surgical planning in patients with CHD^41^. In the setting of AV-valve regurgitation, 3DE directly traces the orifice area without making assumptions on the shape of the vena contracta. Simultaneous orthogonal visualization of different planes allows a more precise depiction of the best cross-sectional cutting plane to trace the vena contracta circumference. In adults, a strong correlation has been proven between the 3D-VCRA and the regurgitation area calculated by MRI^42^. Adubiab et al. showed that measurements of 3D-VCRA were superior to 2DE determination of regurgitation severity^40^. However, experience with quantitative 3DE in children with CHD is still limited, with only a few studies assessing its potential value. Here, we showed that early changes in 3D-VCRA had significantly higher predictive accuracy than both the 2DE qualitative postoperative MR assessment and the early changes of 2D-VCRA.

AV-valve stenosis is also known to be associated with adverse outcomes in patients after repair of CMVD or AV-canal^10, 29, 43^. MV stenosis is generally assessed by 2DE using mean AV-valve inflow gradients; however, this measurement presents several challenges. First, many patients with CHD may have left ventricular systolic or diastolic dysfunction and therefore elevated left ventricular end diastolic pressure (LVEDP), with consequent possible underestimation of the stenosis, if measured using the gradient alone. Additionally, patients may have limited flow across the valve due to the presence of an atrial shunt or low cardiac output. Since gradients are flow dependent, this may also underestimate the magnitude of the stenosis if assessed by mean inflow gradient only. Our study investigated the utility of 3DE assessment in stenotic valves, and found that early changes in 3D-EOA were accurate in predicting MV reoperation. Most importantly, early changes in 3D-EOA had significantly higher prognostic value than early changes in 2D mean gradient.

To improve risk stratification and facilitate potential application of these findings into clinical practice, decision-tree predictive algorithms were developed and subsequently bootstrapped validated. Potentially, these 3DE parameters could be utilized either pre-operatively or in the operating room. Early changes in 3D-EOA and 3D-VCRA measured in the operating room may inform decision to return to bypass, or, if these changes are measured at discharge, they may inform earlier follow-up or early MV reoperation.

### Study limitations

Since this is a retrospective study, loss of information occurred and not all patients had 3DE data available. As this observational study was conducted in a clinical setting with a variety of providers, the criteria for MV reoperation were not defined and surgical details were not investigated. The 3DE parameters identified as independent predictors were calculated using postoperative changes assessed at discharge. We realize the clinical importance of identifying pre-operative predictors or using the bypass echocardiogram to help guide patient management; however, in the future, these measurements may be potentially used in the operating room. Additionally, this study was limited by constraints of imaging, particular image resolution and low frame rate. Despite these limitations, we believe this study provides a valuable framework for future research investigations in the field of 3DE and in the assessment of CMVD.

### Conclusions

In a large cohort of patients with CMVD, 31% of patients required MV-reoperation at a median follow-up time of 5 years. Age <1 year, presence of tethered leaflets, evidence of moderate or greater postoperative MR on 2DE, as well as early changes in the 3D-EOA and 3D-VCRA were independent predictors of MV-reoperation. 3DE parameters were found to have significantly higher predictive value compared to 2DE parameters, suggesting added value of the 3DE assessment. Decision-tree predictive algorithms were developed based on these findings to classify patients in terms of risk stratification and to serve as a support tool for assessment and clinical decision making.

## Data Availability

The authors confirm that the data supporting the findings of this study are available within the article and its supplementary materials.

## Acknowledgements

The authors thank Drs. Alejandra Bueno, Erin Krizman and Patrick Myers for support with data collection. The authors also thank Dr. Jane Newburger for valuable comments on the manuscript.

## Sources of Funding

Nora Lang received the Kaplan Meier Fellowship and a fellowship from the German Research Foundation.

## Disclosures

There are no disclosures.

## NON-STANDARD ABBREVIATIONS AND ACRONYMS

2DE: 2-dimensional echocardiography
3DE: 3-dimensional echocardiography
AUC: area under the curve
CI: confidence interval
EOA: effective orifice area
HLHS: hypoplastic left heart syndrome
HR: hazard ratio
IQR: interquartile range
LVEDP: left ventricular end diastolic pressure
MD: mixed disease
MR: mitralregurgitation
MS: mitral stenosis
MV: mitral valve
N: number
ROC: receiver operating characteristic
VCRA: vena contracta regurgitant area

## Notes

### Competing Interest Statement

The authors have declared no competing interest.

### Author Declarations

The Institutional Review Board at Boston Children's Hospital approved the study with a waiver for informed consent (IRB-P00023266).

## REFERENCES

1. Baird CW, Marx GR, Borisuk M, Emani S, del Nido PJ. Review of Congenital Mitral Valve Stenosis: Analysis, Repair Techniques and Outcomes. Cardiovasc Eng Technol. 2015;6:167–173. doi: 10.1007/s13239-015-0223-0 10.1007/s13239-015-0223-0 [pii]

2. Seguela PE, Houyel L, Acar P. Congenital malformations of the mitral valve. Arch Cardiovasc Dis. 2011;104:465–479. doi: S1875-2136(11)00247-6 [pii] 10.1016/j.acvd.2011.06.004

3. Delmo Walter EM, Hetzer R. Repair for Congenital Mitral Valve Stenosis. Semin Thorac Cardiovasc Surg Pediatr Card Surg Annu. 2018;21:46–57. doi: S1092-9126(17)30002-9 [pii] 10.1053/j.pcsu.2017.11.008

4. van Rijk-Zwikker GL, Delemarre BJ, Huysmans HA. Mitral valve anatomy and morphology: relevance to mitral valve replacement and valve reconstruction. J Card Surg. 1994;9:255–261.

5. Kalfa D, Vergnat M, Ly M, Stos B, Lambert V, Baruteau A, Belli E. A standardized repair-oriented strategy for mitral insufficiency in infants and children: midterm functional outcomes and predictors of adverse events. J Thorac Cardiovasc Surg. 2014;148:1459–1466. doi: S0022-5223(14)00264-5 [pii] 10.1016/j.jtcvs.2014.02.057

6. Delmo Walter EM, Komoda T, Siniawski H, Hetzer R. Surgical reconstruction techniques for mitral valve insufficiency from lesions with restricted leaflet motion in infants and children. J Thorac Cardiovasc Surg. 2012;143:S48–53. doi: S0022-5223(11)01152-4 [pii] 10.1016/j.jtcvs.2011.10.033

7. Oppido G, Davies B, McMullan DM, Cochrane AD, Cheung MM, d’Udekem Y, Brizard CP. Surgical treatment of congenital mitral valve disease: midterm results of a repair-oriented policy. J Thorac Cardiovasc Surg. 2008;135:1313–1320; discussion 1320-1311. doi: S0022- 5223(07)01997-6 [pii] 10.1016/j.jtcvs.2007.09.071

8. Delmo Walter EM, Komoda T, Siniawski H, Miera O, Van Praagh R, Hetzer R. Long term surgical outcome of mitral valve repair in infants and children with Shone’s anomaly. Eur J Cardiothorac Surg. 2013;43:473–481; discussion 481-472. doi: ezs325 [pii] 10.1093/ejcts/ezs325

9. Selamet Tierney ES, Graham DA, McElhinney DB, Trevey S, Freed MD, Colan SD, Geva T. Echocardiographic predictors of mitral stenosis-related death or intervention in infants. Am Heart J. 2008;156:384–390. doi: S0002-8703(08)00229-9 [pii] 10.1016/j.ahj.2008.03.019

10. Sughimoto K, Konstantinov IE, d’Udekem Y, Brink J, Zannino D, Brizard CP. Mid-term outcomes of congenital mitral valve surgery: Shone’s syndrome is a risk factor for death and reintervention. Interact Cardiovasc Thorac Surg. 2017;25:734–739. doi: 4077061 [pii] 10.1093/icvts/ivx211

11. Ashikhmina E, Shook D, Cobey F, Bollen B, Fox J, Liu X, Worthington A, Song P, Shernan S. Three-dimensional versus two-dimensional echocardiographic assessment of functional mitral regurgitation proximal isovelocity surface area. Anesth Analg. 2015;120:534–542. doi: 10.1213/ANE.0000000000000409 00000539-201503000-00011 [pii]

12. Bhave NM, Lang RM. Quantitative echocardiographic assessment of native mitral regurgitation: two and three-dimensional techniques. J Heart Valve Dis. 2011;20:483–492.

13. Cantinotti M, Giordano R, Koestenberger M, Voges I, Santoro G, Franchi E, Assanta N, Valverde I, Simpson J, Kutty S. Echocardiographic examination of mitral valve abnormalities in the paediatric population: current practices. Cardiol Young. 2020;30:1–11. doi: S1047951119003196 [pii] 10.1017/S1047951119003196

14. Colen T, Smallhorn JF. Three-dimensional echocardiography for the assessment of atrioventricular valves in congenital heart disease: past, present and future. Semin Thorac Cardiovasc Surg Pediatr Card Surg Annu. 2015;18:62–71. doi: S1092-9126(15)00006-X [pii] 10.1053/j.pcsu.2015.01.003

15. Bhatt HV, Spivack J, Patel PR, El-Eshmawi A, Amir Y, Adams DH, Fischer GW. Correlation of 2-Dimensional and 3-Dimensional Echocardiographic Analysis to Surgical Measurements of the Tricuspid Valve Annular Diameter. J Cardiothorac Vasc Anesth. 2019;33:137–145. doi: S1053-0770(18)30381-1 [pii] 10.1053/j.jvca.2018.05.048

16. Baldea SM, Velcea AE, Rimbas RC, Andronic A, Matei L, Calin SI, Muraru D, Badano LP, Vinereanu D. 3-D Echocardiography Is Feasible and More Reproducible than 2-D Echocardiography for In-Training Echocardiographers in Follow-up of Patients with Heart Failure with Reduced Ejection Fraction. Ultrasound Med Biol. 2021;47:499–510. doi: S0301- 5629(20)30485-3 [pii] 10.1016/j.ultrasmedbio.2020.10.022

17. Acar P, Laskari C, Rhodes J, Pandian N, Warner K, Marx G. Three-dimensional echocardiographic analysis of valve anatomy as a determinant of mitral regurgitation after surgery for atrioventricular septal defects. Am J Cardiol. 1999;83:745–749. doi: S0002914998009825 [pii] 10.1016/s0002-9149(98)00982-5

18. Takahashi K, Mackie AS, Thompson R, Al-Naami G, Inage A, Rebeyka IM, Ross DB, Khoo NS, Colen T, Smallhorn JF. Quantitative real-time three-dimensional echocardiography provides new insight into the mechanisms of mitral valve regurgitation post-repair of atrioventricular septal defect. J Am Soc Echocardiogr. 2012;25:1231–1244. doi: S0894- 7317(12)00678-5 [pii] 10.1016/j.echo.2012.08.011

19. Takahashi K, Guerra V, Roman KS, Nii M, Redington A, Smallhorn JF. Three dimensional echocardiography improves the understanding of the mechanisms and site of left atrioventricular valve regurgitation in atrioventricular septal defect. J Am Soc Echocardiogr. 2006;19:1502–1510. doi: S0894-7317(06)00719-X [pii] 10.1016/j.echo.2006.07.011

20. Takahashi K, Inage A, Rebeyka IM, Ross DB, Thompson RB, Mackie AS, Smallhorn JF. Real-time 3-dimensional echocardiography provides new insight into mechanisms of tricuspid valve regurgitation in patients with hypoplastic left heart syndrome. Circulation. 2009;120:1091–1098. doi: CIRCULATIONAHA.108.809566 [pii] 10.1161/CIRCULATIONAHA.108.809566

21. Wolbers M, Koller MT, Stel VS, Schaer B, Jager KJ, Leffondre K, Heinze G. Competing risks analyses: objectives and approaches. Eur Heart J. 2014;35:2936–2941. doi: ehu131 [pii] 10.1093/eurheartj/ehu131

22. Staffa SJ, Zurakowski D. Competing risks analysis of time-to-event data for cardiovascular surgeons. J Thorac Cardiovasc Surg. 2020;159:2459–2466 e2455. doi: S0022- 5223(19)32777-1 [pii] 10.1016/j.jtcvs.2019.10.153

23. Zhou B FJ, Laid G. . Stat Med. 2013 ;32 :3804-3811. Goodness-of-fit test for proportional subdistribution hazards model. Stat Med 2013 ;32 :3804–3811 2013.

24. DeLong ER, DeLong DM, Clarke-Pearson DL. Comparing the areas under two or more correlated receiver operating characteristic curves: a nonparametric approach. Biometrics. 1988;44:837–845.

25. Staffa SJ, Zurakowski D. Statistical Development and Validation of Clinical Prediction Models. Anesthesiology. 2021;135:396–405. doi: 116300 [pii] 10.1097/ALN.0000000000003871

26. Baghaei R, Tabib A, Jalili F, Totonchi Z, Mahdavi M, Ghadrdoost B. Early and Mid Term Outcome of Pediatric Congenital Mitral Valve Surgery. Res Cardiovasc Med. 2015;4:e28724. doi: 10.5812/cardiovascmed.28724v2

27. Stephens EH, Han J, Ginns J, Rosenbaum M, Chai P, Bacha E, Kalfa D. Outcomes and Prognostic Factors for Adult Patients With Congenital Heart Disease Undergoing Primary or Reoperative Systemic Atrioventricular Valve Surgery. World J Pediatr Congenit Heart Surg. 2017;8:346–353. doi: 10.1177/2150135117692974

28. Schleiger A, Miera O, Peters B, Schmitt KRL, Kramer P, Buracionok J, Murin P, Cho MY, Photiadis J, Berger F, et al. Long-term results after surgical repair of atrioventricular septal defect. Interact Cardiovasc Thorac Surg. 2019;28:789–796. doi: 5258054 [pii] 10.1093/icvts/ivy334

29. Ijsselhof R, Gauvreau K, Nido PD, Nathan M. Atrioventricular Valve Function Predicts Reintervention in Complete Atrioventricular Septal Defect. World J Pediatr Congenit Heart Surg. 2020;11:247–248. doi: 10.1177_2150135119893648 [pii] 10.1177/2150135119893648

30. Fong LS, Betts K, Ayer J, Andrews D, Nicholson IA, Winlaw DS, Orr Y. Predictors of reoperation and mortality after complete atrioventricular septal defect repair. Eur J Cardiothorac Surg. 2021;61:45–53. doi: 6277380 [pii] 10.1093/ejcts/ezab221

31. Prakash A, Lacro RV, Sleeper LA, Minich LL, Colan SD, McCrindle B, Covitz W, Golding F, Hlavacek AM, Levine JC, et al. Challenges in echocardiographic assessment of mitral regurgitation in children after repair of atrioventricular septal defect. Pediatr Cardiol. 2012;33:205–214. doi: 10.1007/s00246-011-0107-5

32. Yosefy C, Hung J, Chua S, Vaturi M, Ton-Nu TT, Handschumacher MD, Levine RA. Direct measurement of vena contracta area by real-time 3-dimensional echocardiography for assessing severity of mitral regurgitation. Am J Cardiol. 2009;104:978–983. doi: S0002- 9149(09)01123-0 [pii] 10.1016/j.amjcard.2009.05.043

33. Kaza E, Marx GR, Kaza AK, Colan SD, Loyola H, Perrin DP, Del Nido PJ. Changes in left atrioventricular valve geometry after surgical repair of complete atrioventricular canal. J Thorac Cardiovasc Surg. 2012;143:1117–1124. doi: S0022-5223(11)01132-9 [pii] 10.1016/j.jtcvs.2011.06.044

34. Sugeng L, Weinert L, Lang RM. Real-time 3-dimensional color Doppler flow of mitral and tricuspid regurgitation: feasibility and initial quantitative comparison with 2-dimensional methods. J Am Soc Echocardiogr. 2007;20:1050–1057. doi: S0894-7317(07)00120-4 [pii] 10.1016/j.echo.2007.01.032

35. Zeng X, Levine RA, Hua L, Morris EL, Kang Y, Flaherty M, Morgan NV, Hung J. Diagnostic value of vena contracta area in the quantification of mitral regurgitation severity by color Doppler 3D echocardiography. Circ Cardiovasc Imaging. 2011;4:506–513. doi: CIRCIMAGING.110.961649 [pii] 10.1161/CIRCIMAGING.110.961649

36. Thavendiranathan P, Liu S, Datta S, Rajagopalan S, Ryan T, Igo SR, Jackson MS, Little SH, De Michelis N, Vannan MA. Quantification of chronic functional mitral regurgitation by automated 3-dimensional peak and integrated proximal isovelocity surface area and stroke volume techniques using real-time 3-dimensional volume color Doppler echocardiography: in vitro and clinical validation. Circ Cardiovasc Imaging. 2013;6:125–133. doi: CIRCIMAGING.112.980383 [pii] 10.1161/CIRCIMAGING.112.980383

37. Marsan NA, Westenberg JJ, Ypenburg C, Delgado V, van Bommel RJ, Roes SD, Nucifora G, van der Geest RJ, de Roos A, Reiber JC, et al. Quantification of functional mitral regurgitation by real-time 3D echocardiography: comparison with 3D velocity-encoded cardiac magnetic resonance. JACC Cardiovasc Imaging. 2009;2:1245–1252. doi: S1936-878X(09)00597- X [pii] 10.1016/j.jcmg.2009.07.006

38. Zamorano J, de Agustin JA. Three-dimensional echocardiography for assessment of mitral valve stenosis. Curr Opin Cardiol. 2009;24:415–419. doi: 10.1097/HCO.0b013e32832e165b

39. Medvedofsky D, Maffessanti F, Weinert L, Tehrani DM, Narang A, Addetia K, Mediratta A, Besser SA, Maor E, Patel AR, et al. 2D and 3D Echocardiography-Derived Indices of Left Ventricular Function and Shape: Relationship With Mortality. JACC Cardiovasc Imaging. 2018;11:1569–1579. doi: S1936-878X(17)30904-X [pii] 10.1016/j.jcmg.2017.08.023

40. Abudiab MM, Chao CJ, Liu S, Naqvi TZ. Quantitation of valve regurgitation severity by three-dimensional vena contracta area is superior to flow convergence method of quantitation on transesophageal echocardiography. Echocardiography. 2017;34:992–1001. doi: 10.1111/echo.13549

41. Simpson J, Lopez L, Acar P, Friedberg M, Khoo N, Ko H, Marek J, Marx G, McGhie J, Meijboom F, et al. Three-dimensional echocardiography in congenital heart disease: an expert consensus document from the European Association of Cardiovascular Imaging and the American Society of Echocardiography. Eur Heart J Cardiovasc Imaging. 2016;17:1071–1097. doi: jew172 [pii] 10.1093/ehjci/jew172

42. Maragiannis D, Little SH. 3D vena contracta area to quantify severity of mitral regurgitation: a practical new tool? Hellenic J Cardiol. 2013;54:448–454.

43. Stellin G, Padalino MA, Vida VL, Boccuzzo G, Orru E, Biffanti R, Milanesi O, Mazzucco A. Surgical repair of congenital mitral valve malformations in infancy and childhood: a single-center 36-year experience. J Thorac Cardiovasc Surg. 2010;140:1238–1244. doi: S0022- 5223(10)00484-8 [pii] 10.1016/j.jtcvs.2010.05.016

